# Quantitative detection of SARS-CoV-2 B.1.1.7 variant in wastewater by allele-specific RT-qPCR

**DOI:** 10.1101/2021.03.28.21254404

**Authors:** Wei Lin Lee, Kyle A McElroy, Federica Armas, Maxim Imakaev, Xiaoqiong Gu, Claire Duvallet, Franciscus Chandra, Hongjie Chen, Mats Leifels, Samuel Mendola, Róisín Floyd-O’Sullivan, Morgan M Powell, Shane T Wilson, Fuqing Wu, Amy Xiao, Katya Moniz, Newsha Ghaeli, Mariana Matus, Janelle Thompson, Eric J Alm

## Abstract

Wastewater-based epidemiology (WBE) has emerged as a critical public health tool in tracking the SARS-CoV-2 epidemic. Monitoring SARS-CoV-2 variants of concern in wastewater has to-date relied on genomic sequencing, which lacks sensitivity necessary to detect low variant abundances in diluted and mixed wastewater samples. Here, we develop and present an open-source method based on allele specific RT-qPCR (AS RT-qPCR) that detects and quantifies the B.1.1.7 variant, targeting spike protein mutations at three independent genomic loci highly predictive of B.1.1.7 (HV69/70del, Y144del, and A570D). Our assays can reliably detect and quantify low levels of B.1.1.7 with low cross-reactivity, and at variant proportions between 0.1% and 1% in a background of mixed SARS-CoV-2. Applying our method to wastewater samples from the United States, we track B.1.1.7 occurrence over time in 19 communities. AS RT-qPCR results align with clinical trends, and summation of B.1.1.7 and wild-type sequences quantified by our assays strongly correlate with SARS-CoV-2 levels indicated by the US CDC N1/N2 assay. This work paves the path for rapid inexpensive surveillance of B.1.1.7 and other SARS-CoV-2 variants in wastewater.

## INTRODUCTION

The COVID-19 pandemic has caused an unprecedented disruption in all aspects of life globally. Recent rollout of vaccines across a number of countries has been an important milestone in fighting the pandemic. However, the emergence of new variants of concern (VOCs) such as B.1.1.7 and B.1.351 suggests that continued vigilance is required to control the COVID-19 pandemic (Mascola, Graham, & Fauci, 2021). For example, the B.1.1.7 variant contains spike protein mutations D614G and N501Y that have been reported to increase transmission and to subsequently cause a more severe COVID-19 manifestation with increased hospitalization and mortality rates (Davies et al., 2021; Hou et al., 2020; Korber et al., 2020; Rambaut et al., 2020). Moreover, both the B.1.1.7 and B.1.351 variants contain extensive mutations in the spike protein that cause increased resistance to neutralization by antibodies derived from convalescent patients’ plasma and vacinee sera (Wang et al., 2021). The potential public health challenges posed by these emerging VOCs warrant extensive real-time surveillance at the community level to inform efforts in controlling the pandemic (CDC, 2020).

While the gold standard for disease surveillance is through clinically derived data (*i*.*e*. swabbing of the upper respiratory tract followed by qPCR), wastewater-based epidemiology (WBE) has emerged as a cost-effective and non-invasive complement to clinical testing during this COVID-19 pandemic (Hart & Halden, 2020). WBE can provide real-time, unbiased disease surveillance at the community level due to the fact that wastewater contains aggregated population health data (Polo et al., 2020; Thompson, 2020). Numerous countries such as the USA (Wu et al., 2020), the Netherlands (Medema et al., 2020), Australia (Ahmed et al., 2020), and Spain (Randazzo et al., 2020) have reported the presence of SARS-CoV 2 in their wastewater. WBE is actively utilized and supported by government and public health entities: national programs have been established in the US (NWSS, 2021), and the European Commission recently recommended WBE as an approach for the systematic surveillance of SARS-CoV-2 and its variants in the EU (EU Commission, 2021).

In light of new emerging VOCs, efforts have been made to utilize WBE in tracking SARS-CoV 2 variants that are circulating in the community. However, the most common method applied to wastewater is currently next generation sequencing (NGS), either by sequencing the viral metagenome of wastewater (Crits-Christoph et al., 2020) or by amplifying and then sequencing SARS-CoV-2 genome extracted from wastewater samples (Prado et al., 2021). While these early results are promising, many report inconsistent and/or low sequencing coverage, likely due to the low input concentrations of SARS-CoV-2 in wastewater (Fontenele et al., 2021; Jahn et al., 2021; Prado et al., 2021). Therefore, NGS-based approaches for WBE will require considerable additional optimization before they can be robustly applied to a broad variety of wastewater samples and to generate reliable high-quality data that can be used to inform public health responses. Furthermore, universally implementing NGS-based wastewater surveillance will require additional specialised infrastructure and standardized protocols (Gwinn, MacCannell, & Armstrong, 2019). As such, there is a critical need for a simple, accurate and convenient diagnostic assay for the identification of SARS-CoV-2 variants which is suited for wastewater surveillance and which can leverage existing experimental pipelines and infrastructure. To address this gap, we developed and validated a method that is based upon RT-qPCR, a widely applied and commonly available laboratory assay used in diagnostic testing for SARS-CoV-2 (Lu et al., 2020) and broadscale WBE efforts (Wu et al., 2021). This method consists of a panel of allele specific (AS) RT-qPCR-based assays which we apply to identify and quantify the B.1.1.7 variant of SARS-CoV-2 and which can be easily expanded to target additional variants of concern.

AS-PCR was developed over 30 years ago (Petruska et al., 1988; Wu et al., 1989) and is commonly used for molecular genotyping in laboratory diagnostics (Coleman & Tsongalis, 2007). Here, we leverage AS-PCR to detect mutations in the SARS-CoV-2 genome in wastewater, with application to VOC tracking. AS-PCR permits the direct detection of one or more mutations in the genome of an organism or interest. It involves two parallel reactions, each including a wild-type (WT) or mutation specific forward primer and a common reverse primer (or vice versa), in which the WT or mutation specific primer is designed with its 3′ end complementary to each mutation. Introduction of a synthetic mismatch near the 3’ terminal end of both WT and mutant primers improves specificity by significantly destabilizing hybridization of non-target sequences (Newton et al, 1991). The 3′ mismatch between the oligonucleotide primer and the genome template is refractory to primer extension by *Thermus aquaticus* (Taq) polymerase, conferring up to single nucleotide discrimination. Further AS-PCR can be performed with fluorescent dyes quantitatively (as AS-qPCR) to visualize product accumulation with hydrolysis probes, of which quantification cycle values (Ct) enable accurate quantitative measurements. When coupled with reverse transcription (RT) as is required for RNA viruses such as SARS-CoV-2, the method is referred to as AS RT-qPCR.

Our AS RT-qPCR panel for SARS-CoV-2 wastewater-based variant tracking consists of three pairs of primer-probe sets that target B.1.1.7-specific spike protein mutations HV69/70del, Y144del, and A570D. These three pairs of primer-probe sets enable the detection and discrimination of SARS-CoV-2 variant B.1.1.7 sequence from the WT sequence. The assay presented here uses identical reagents, instruments and workflow as existing SARS-CoV-2 RT-qPCR diagnostic assays, thus potentially allowing for its immediate implementation on a global scale. We performed analytical assessments of the primer sets to confirm that they have amplification efficiencies above 90%, possess similar sensitivities to the most commonly used and CDC recommended N1 and N2 assays, and reliably enable specific detection and accurate quantitation in a background of RNA of the opposite genotype (B.1.1.7 in WT and vice versa). We also applied this panel of assays for quantitative detection of B.1.1.7 in SARS-CoV-2 positive wastewater samples. This work demonstrates the utility of AS RT-qPCR for variant detection in wastewater, and could be readily applied to track the spread of B.1.1.7 and adapted to track other VOCs through wastewater surveillance.

## RESULTS

### AS RT-qPCR primer design strategy

Our AS RT-qPCR assay is designed to enable quantitation of the B.1.1.7 variant and WT SARS-CoV-2. We focused on the loci that can distinguish B.1.1.7 from other variants. For each locus, we designed two allele-specific (AS) forward primers, one for the WT allele and one for the mutant allele, with the 3’ end of these primers terminating at the mutation to be assayed. Parallel quantitative reverse transcription–PCR (RT-qPCR) reactions targeting both the WT and variant sequence are required to determine the variant frequency at the corresponding locus. Each of these contain one of the allele-specific (AS) forward primers, a common reverse primer and a common probe. Nucleotide discrimination of the AS primers employs the principle of thermodynamic instability of the 3’ end being refractory to polymerase extension. To further increase thermodynamic instability for enhanced template discrimination, a synthetic mismatch near the final 3’ nucleotide has been incorporated. Using this strategy, we designed AS RT-qPCR primers targeting spike protein mutations characteristic of B.1.1.7 that would enable discrimination between B.1.1.7 and WT sequences.

We screened these primers against B.1.1.7 and WT synthetic RNA to identify combinations that could be utilized to discriminate between B.1.1.7 and WT. This initial screen identified primers targeting HV69-70del, Y144del and A570D as having the greatest sensitivity (lowest cycle threshold) and specificity (highest difference in cycle thresholds between specific and off target amplification). In this work we report detailed analytical validation of the sensitivity and specificity of these three pairs of primer-probe sets. The schematic of our allele-specific RT-qPCR for determination of variant frequencies is illustrated in **Figure 1**.

**Figure 1.**
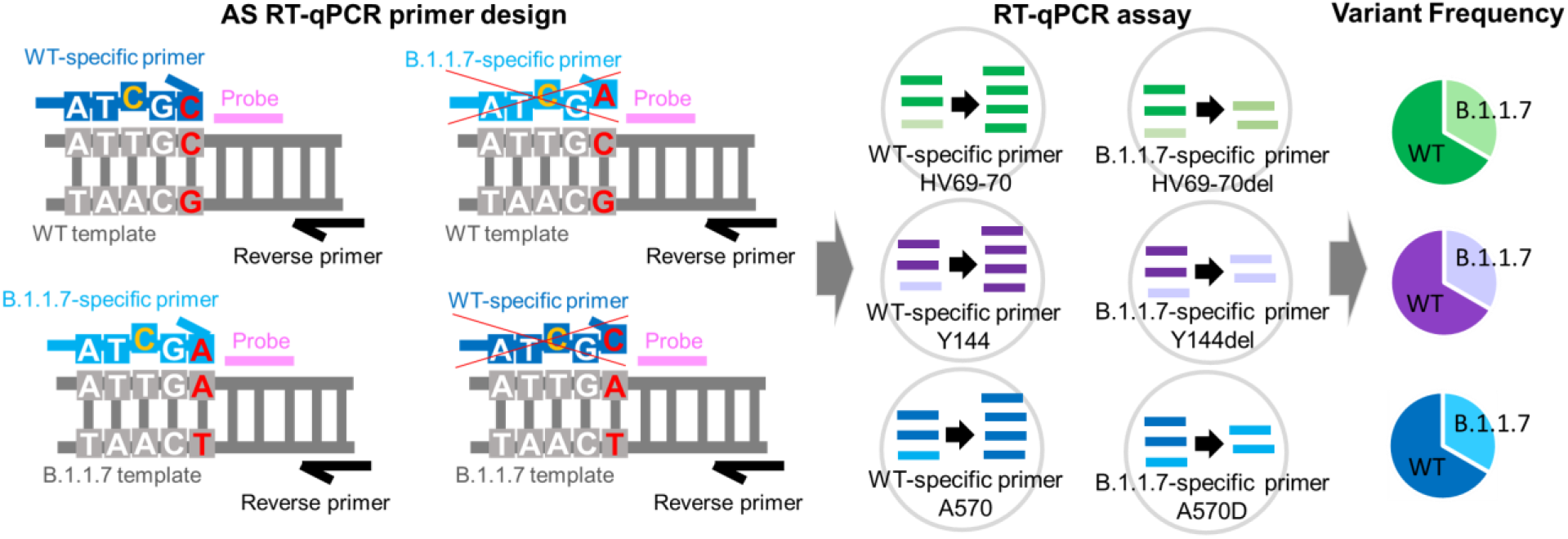
Schematic of our panel of three AS RT-qPCR assays. (Left panel) Each assay requires a common reverse primer (black) and probe (pink), paired with a WT or mutation-specific forward primer (light and dark blue). This allele-specific primer is designed to target the mutation of interest (red) at the 3′ end of the mutation-specific primer and contains a synthetic mismatch (orange) near the 3’ end to enhance assay specificity. Sequences shown represent WT and B.1.1.7 primer and target sequences for loci A570D. (Middle panel) Two individual RT-qPCR reactions are needed to assay for variant frequencies at each locus. (Right panel) Variant frequencies can be calculated after converting threshold cycles to genome copies using their respective standard curves.

### Predictive value of SARS-CoV-2 spike protein variants targeted for B.1.1.7 detection

We looked at how predictive our selected mutations HV69-70del, Y144del and A570D were for detection of the B.1.1.7 lineage (**Table 1**). We downloaded all available genomes in GISAID as of 11 Mar 2021 and aligned them to the SARS-CoV-2 reference genome (NC_045512.2) to calculate the percent of B.1.1.7 and non-B.1.1.7 genomes containing each target mutation. HV69-70del is present in 98.7% of sequences in the B.1.1.7 lineage but also present in 2.5% of other lineages. Deletion Y144del is present in 97.9% of publicly available B.1.1.7 sequences and only in 0.5% WT and non-B.1.1.7 genomes. A570D is conferred by a C to A mutation at nucleotide 23,271. 99.9% of sequences in the B.1.1.7 lineage show this mutation, while it has been reported in only 0.02% of WT and non-B.1.1.7 sequences. Given the high specificity of these mutations to B.1.1.7, the co-occurrence of these three mutations in wastewater is likely highly indicative of SARS-CoV-2 RNA with B.1.1.7 lineage.

**Table 1.** Frequency of targeted mutations in SARS-CoV-2 B.1.1.7 and non-B.1.1.7 strains (including WT and other non-B.1.1.7 variants); Sequences obtained from GISAID database as of 11 Mar 2021.

| Amino acid position(s) of targeted mutations | S gene nucleotide changes | Frequency in B.1.1.7 | Frequency in other variants |
| --- | --- | --- | --- |
| HV69-70del | Deletion of 21,765-21,770 | Present in 98.7% of B.1.1.7 | Present in 2.5% of WT and non-B.1.1.7 variants |
| Y144del | Deletion of 21,991-21,993 | Present in 97.9% of B.1.1.7 | Present in 0.5% of WT and non-B.1.1.7 variants |
| A570D | C23271A | Present in 99.9% of B.1.1.7 | Present in 0.02% of WT and non-B.1.1.7 variants |

### Specificity of AS RT-qPCR primers for their respective RNA

We examined the specificity of the three designed AS RT-qPCR assays for their respective WT and mutant genome targets in the SARS-CoV-2 RNA (HV69-70 del, Y144del and A570D) by screening them against full length synthetic RNA constructs of the B.1.1.7 and WT genotypes (**Figure 2**). The amplification efficiency of each primer and probe set were between 90.6 to 105% for the correct RNA (*i*.*e*. B.1.1.7 assay for B.1.1.7 RNA, and WT assay for WT RNA). Next, we next determined the levels of cross reactivity for RNA of the opposite genotype by testing each combination of WT and mutant primers. Cross reactivity did not occur for WT HV69-70 and WT Y144 primers, against up to 10^5^ and 10^4^ copies of B.1.1.7 RNA respectively (**Figure 2**, top left panel). WT A570 primers were less specific but still strongly discriminant against the B.1.1.7 variant sequence **(Figure 2**; left column, bottom panel). Cross-reactivity for all three B.1.1.7-specific primers was minimal, observed only for 10^4^ and above copies of WT RNA per µL (**Figure 2**, right column) which suggests that the assay can detect low B.1.1.7 variant frequencies in the presence of a strong background of WT sequences.

**Figure 2.**
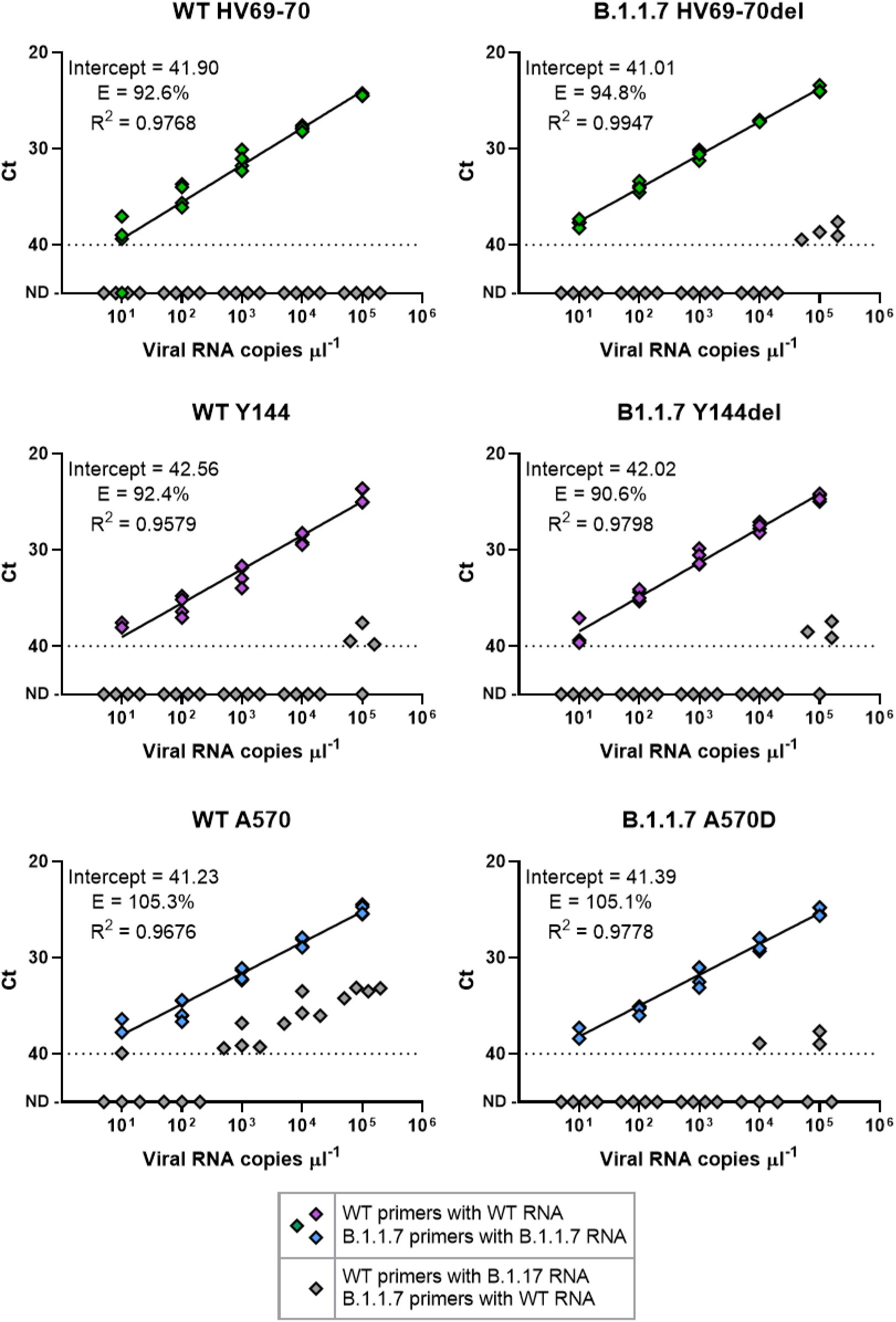
Specificity and cross-reactivity of AS RT-qPCR primers against WT and B.1.1.7 RNA in nuclease-free water. PCR efficiency and y-intercept cycle threshold (Ct) values were calculated for each of six primer–probe sets against tenfold dilutions of synthetic full-length SARS-CoV-2 RNA. Data shown reflect two sets of independent measurements taken on different days. Green, purple and blue diamonds represent tests against the matching genotype (WT-specific primers to WT RNA and B.1.1.7-specific primers to B.1.1.7 RNA) and grey diamonds denote tests against RNA of the opposite genotype.

### Comparison of Ct values of AS RT-qPCR primers to US CDC N1 and N2 SARS-CoV-2 primers

Given the importance of high sensitivity for effective SARS-CoV-2 detection in an environmental matrix as complex as sewage, we examined the cycle thresholds (Ct) of all three pairs of primer-probe sets in comparison to the N1 and N2 primer-probe sets recommended by the US CDC (**Figure 3**). Having similar or lower Ct values for the same input RNA can be considered a proxy for the overall sensitivity of the assay. We found these three AS RT-qPCR primer-probe sets to have comparable Ct values to N1 and N2 assays for the same amount of input RNA across a 100-fold difference in target RNA concentration. Comparing these results to the y-intercept Ct values in **Figure 2** also supports the conclusion that sensitivities presented here were similar among most of the primer–probe sets, though Y144del may be slightly lower than the other primer-probe sets (higher Ct values in **Figure 3**, purple circles on right panel).

**Figure 3.**
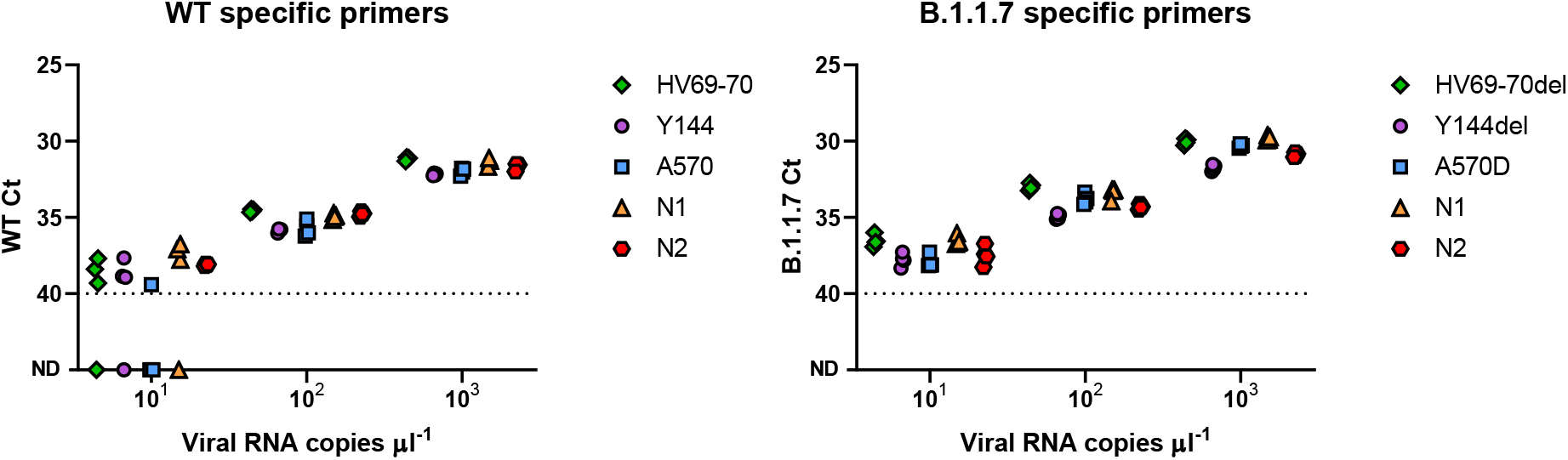
Ct values for the AS RT-qPCR primers in comparison to US CDC N1 and N2 assays. Comparison of Ct values for low-copy number detection of WT and VOC targets suggests similar sensitivity of AS RT-qPCR primers in comparison to US CDC N1 and N2 SARS-CoV-2 primers. Primer-probe sets were tested with tenfold dilutions of their respective full length synthetic SARS-CoV-2 RNA in nuclease-free water. Data shown reflect two sets of independent measurements taken on different days.

### Detection in a background of 10^4^ copies of RNA of the opposite genotype

Due to their aggregated and heterogeneous nature, wastewater samples likely contain a mixture of numerous SARS-CoV-2 variant genotypes which can occur at highly skewed ratios. We looked at how suitable our primer-probe sets were to detect and quantify VOCs in the presence of an abundance of the various genotypes. We examined the cycle thresholds (Ct) of primer-probe sets against their respective full length RNA in the presence or absence of up to 10^4^ copies of RNA of the opposite genotype (**Figure 4**), which constitutes a variant frequency of 0.1%, 1% and 10%, respectively.

**Figure 4.**
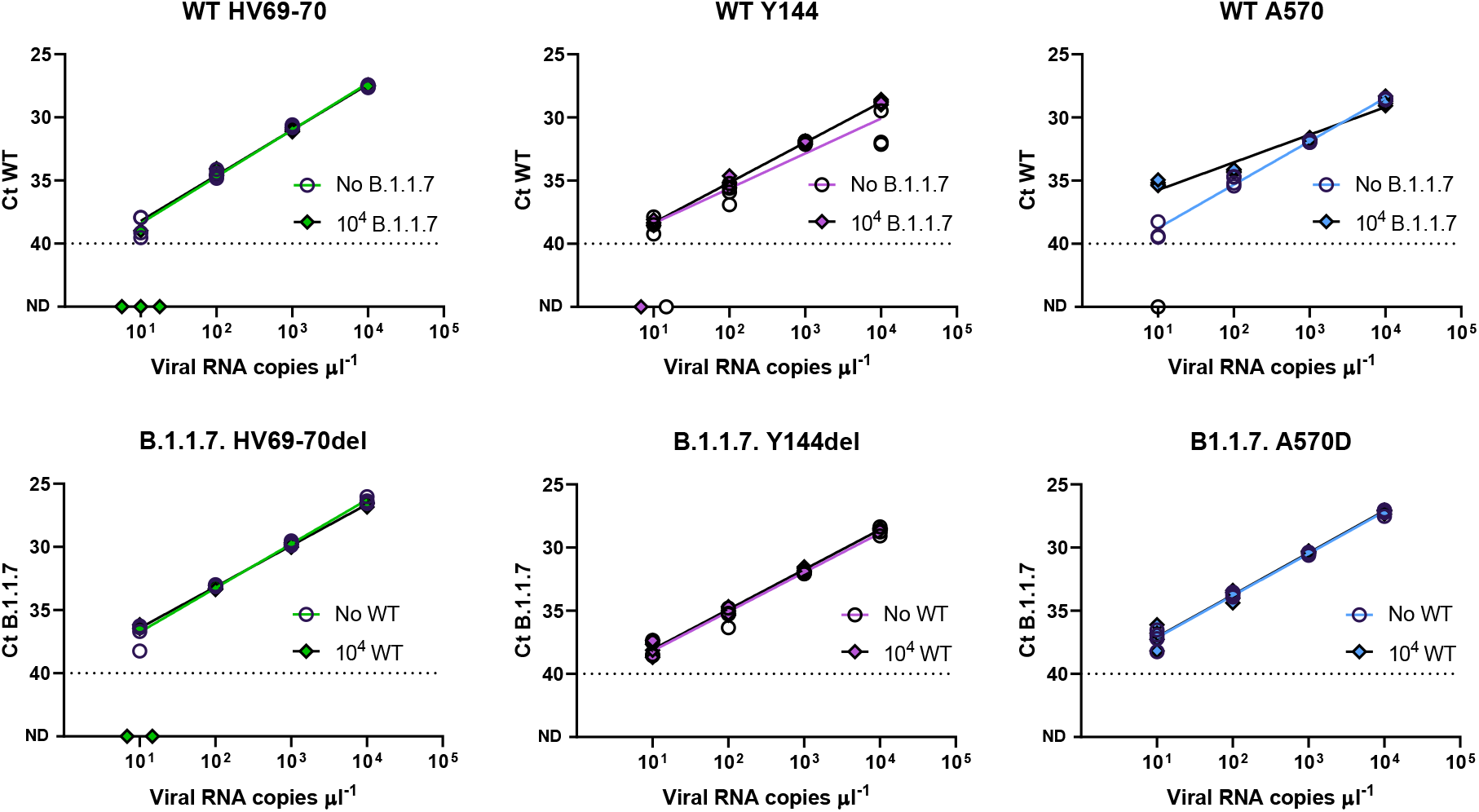
Detection and quantification of synthetic SARS-CoV-2 RNA in the presence of 10^4^ copies of RNA of the opposite genotype. Primer-probe sets were tested with tenfold dilutions of their respective full length synthetic SARS-CoV-2 RNA (x-axis), with and without the presence of 10^4^ copies of RNA of the opposite genotype. Circles and colored lines denote assays performed in the absence of RNA of the opposite genotype. Diamonds and black lines denote assays performed in the presence of 10^4^ copies of RNA of the opposite genotype. Data shown reflect two sets of independent measurements taken on different days.

Overall, the B.1.1.7 primer-probe sets were unaffected by large quantities of WT (background) RNA and the WT primer-probe sets were also largely unaffected by B.1.1.7 background. All of the primer-probe sets presented here were able to detect B.1.1.7 down to a ratio of 1% to WT RNA and vice versa, with cycle thresholds largely unaffected by the presence of 10^4^ copies of RNA of the opposite genotype (**Figure 4**). These results were similarly sensitive down to ratios of 0.1% WT/B.1.1.7 RNA, though sporadic non-detections occurred for WT HV69-70, WT Y144 and B.1.1.7 HV69-70del primers in the presence of RNA of the opposite genotype. WT 144 and WT A570 primers occasionally did not detect 10 copies of their target in the absence of the opposite genotype which is determined to be consistent with their lower overall sensitivities described earlier (**Figure 3**). The majority of primer-probes presented here were unaffected by the presence of 10^4^ copies of RNA of the opposite genotype, except for WT A570 which yielded higher concentrations (lower Ct values) in the presence of high B.1.1.7 background with 10 copies of WT input. Specificity and cross reactivity tests on WT A570 presented earlier in this paper suggest cross-reactivity with the B.1.1.7 VOC resulting in its slight overestimation. Therefore, we have determined that the presented AS RT-qPCR assays are capable of robustly and reproducibly detecting variants of concern in complex matrices in abundances as low as 1%.

### Application on SARS-CoV-2 positive wastewater samples

We next validated the performance of our assay on wastewater samples. We tested samples collected from 16 urban and rural wastewater treatment plants and 3 buildings across 11 US states. For each of these sampling locations, we analyzed samples collected during three distinct time periods: from Fall 2020 (October 20 - November 5, 2020), when no B.1.1.7 was reported in clinical samples in the US; from January 2021 (January 20 - 29, 2021) when B.1.1.7 was first emerging in the US; and between February 24 and March 8, 2021, when B.1.1.7 was circulating in most of the US. Overall, we tested 58 samples, all but two locations had one sample per each time period; MA-2 had two samples tested in each of January 2021 and Fall 2020 periods and FL-6 had no samples in Fall 2020. All of the samples included in this experiment tested positive for SARS-CoV-2 with RT-qPCR using the US CDC N1/N2 assays (**Figure 5**). We also estimated the fraction of B.1.1.7 as a ratio of B.1.1.7 SARS-CoV-2 concentration to the sum of B.1.1.7 and WT concentrations (**Figure 5**).

**Figure 5.**
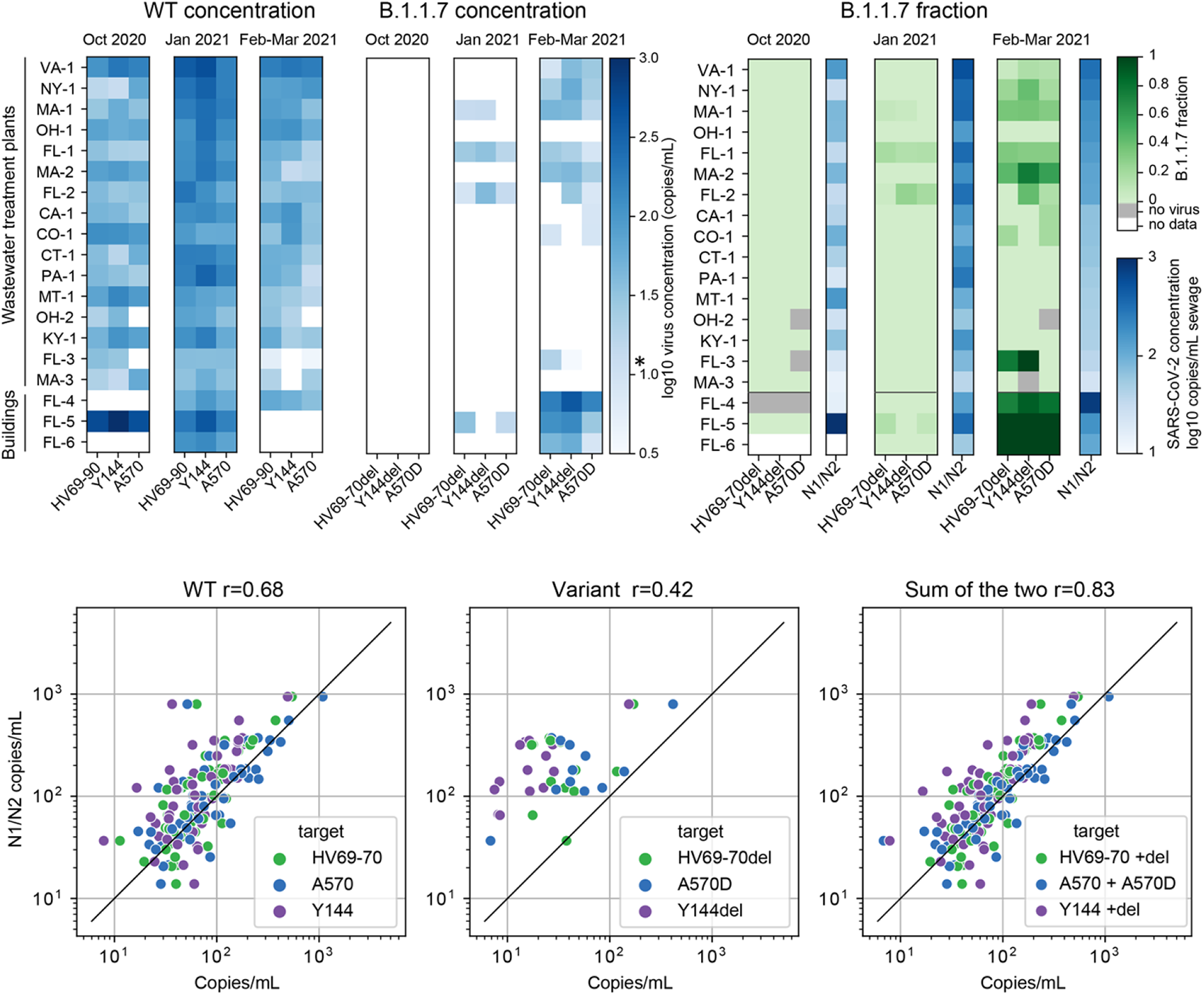
Application of the AS RT-qPCR panel on 16 wastewater treatment plant samples and 3 building wastewaters collected from across the US. (**Top**) Detection and quantitation of WT and B.1.1.7 RNA in wastewater samples collected in the US from October 2020 to March 2021. Locations are ordered from maximum (top) to minimum (bottom) N1/N2 concentration for the most recent (Feb/March 2021) samples. The left plot (blue) shows wastewater concentrations for WT and B.1.1.7 loci. The right plot (green) shows a relative fraction of B.1.1.7 to the total SARS-CoV-2 viral titer derived from the individual pairs of AS RT-qPCR primers. The limit of quantitation (LOQ) for AS RT-qPCR as implemented for these samples is indicated on the heat map scale bar by an asterisk and is higher than for the N1/N2 assay due to sample volume limitation. (**Bottom**) Comparison between N1/N2 concentrations from and concentrations measured by the AS RT-qPCR assay. Each point represents one measurement (each sample thus has three measurements, one per target locus). Left panel compares N1/N2 with the WT loci; middle compares with the B.1.1.7 loci; and right panel compares N1/N2 concentrations with the sum of WT and B.1.1.7 (sum of the total copies, divided by input volume).

The WT sequence was detected in 55 out of 58 samples (for at least 2 target loci), indicating that our WT assays have comparable sensitivity as the routinely used N1/N2 RT-qPCR SARS-CoV-2 protocol. At least two loci associated with B.1.1.7 were detected in 15 out of 56 samples, with 11 additional samples showing positive signals for one mutant locus. B.1.1.7 targets were not detected in the October 2020 samples, reflecting the absence of clinically-confirmed B.1.1.7 in the US at that time. In January 2021, 4 samples were positive in at least 2 target B.1.1.7 loci. Three of these B.1.1.7-positive samples were from Florida, which agrees with the clinical trend that Florida was the US state with the highest fraction of B.1.1.7 cases in January 2021. For comparison, 19.1% of GISAID sequences in Florida in January 2021 were of the B.1.1.7 lineage, compared to 2.8% nation-wide.

In February/March 2021, 12 samples were positive for B.1.1.7 targets. The fraction of the B.1.1.7 variant increased more than 3-fold in all locations between January and February/March 2021. The average fraction of B.1.1.7 for all non-building locations increased from 2.6% to 19.2%. These results are consistent with the average fraction of B.1.1.7 cases in the US according to GISAID for these time periods (4.2% and 13.3% respectively). Interestingly, one building-level sample was found to contain exclusively B.1.1.7 in February-March 2021. This is consistent with our expectation, as for an individual building we could expect only one strain to be circulating in a small number of people.

Finally, we compared results of the AS RT-qPCR assay with the SARS-CoV-2 concentrations measured using US CDC N1 and N2 assays. We found that the concentrations of WT and B.1.1.7 SARS-CoV-2 sequences at the AS RT-qPCR loci were each positively correlated (p<0.001) with SARS-CoV-2 RNA concentrations obtained by the N1/N2 assay. Gene copies per mL of the WT sequences at each locus agreed well with measurements obtained from the N1/N2 assay (**Figure 5, Bottom panel**). The strongest correlations and best match to the N1/N2 data were obtained by summing the concentrations of WT and B.1.1.7 SARS-CoV-2 sequences at the AS RT-qPCR loci, indicating quantitative detection of SARS-CoV-2 RNA at these loci and consistent with the fact that CDC N1/N2 assay does not discriminate between B.1.1.7 and other genotypes.

## DISCUSSION

The present study describes the development and validation of a qPCR-based assay that allows for the detection and quantification of B.1.1.7 in wastewater samples and which is readily adaptable to additional variants of concern. The assay leverages allele specific qPCR (AS-qPCR) and consists of three primer-probe sets that target the HV69-70del, Y144del and A570D spike protein mutations that are specific to B.1.1.7. The primer-probe sets have comparable efficiencies and sensitivities to the widely used US CDC N1 and N2 qPCR assays and can robustly, reproducibly and specifically detect B.1.1.7 in complex matrices and at concentrations between 0.1% and 1% in a background of 10^4^ copies of WT RNA, thus making them suitable for use in wastewater samples with mixed genotypes. Importantly, the primer-probe sets presented here show only minimal cross-reactivity with the wildtype and are capable of quantifying B.1.1.7 RNA at concentrations typically seen in raw wastewater samples. Finally, we apply our method to real wastewater samples and show that it produces interpretable and sensible results. This AS RT-qPCR panel can be performed as six individual one step RT-qPCR reactions, with a similar workflow to existing RT–qPCR assays used by clinical, research and public health laboratories for SARS-CoV-2 diagnostics and wastewater surveillance. This allows for the cost effective and convenient identification and quantitation of B.1.1.7., which can be rapidly integrated into existing wastewater surveillance procedures.

Wastewater surveillance has been increasingly utilized to non-invasively monitor for population-level infections and allows for the tracking of clusters, hotspots and infection trends within a community (Wu et al., 2020; Larsen & Wigginton, 2020). Wastewater surveillance effectively complements clinical testing and is unaffected by biases in clinical data which may be due to limits in testing capacity, the presence of disease symptoms, or treatment seeking behavior. Despite the potential utility of wastewater surveillance for understanding SARS-CoV-2 variant frequencies and dynamics within a community, there exists methodological challenges to variant detection in wastewater. Due to high dilution, wastewater samples contain significantly lower SARS-CoV-2 titers than clinical samples, thus requiring highly sensitive assays. Wastewater is also a complex and composite matrix made up of numerous potential inhibitors, enteric pathogens and - most importantly - multiple viral variants, thus necessitating methods that are highly specific and quantitative. The AS RT-qPCR panel we describe here enables detection and quantitation of B.1.1.7 in wastewater, while using a similar workflow as conventional SARS-CoV-2 RT-qPCR testing. These open source assays thus enable wastewater monitoring programs to identify the occurrence of B.1.1.7, and quantify variant-specific SARS-CoV-2 trends and dynamics at a community level. Most importantly, this AS RT-qPCR-based method is highly modifiable and can be easily extended and adapted to target other variants of concern as soon as they are identified by clinical genomic sequencing surveillance efforts.

The critical need for surveillance of SARS-CoV-2 variants of concern (VOC) has prompted the development of other methods that can track VOCs in wastewater. To date, the most common methods used in frontline laboratories involve sequencing of the entire SARS-CoV-2 genome or targeted sequencing of specific loci of interest, both of which are time consuming, expensive, and face unique data interpretation challenges in wastewater. A qPCR-based approach, on the other hand, is better suited for targeted detection at low viral concentrations, widely available at relatively low cost in laboratories conducting clinical diagnostics, and provides quantitative data for determination of variant abundances. As such, several groups have reported RT-qPCR primer-probe sets for detection of VOCs in wastewater. Most of these groups developed assays targeting only a single site for detection of each VOC lineage. They reported substantially lower sensitivity for their assays in comparison to the US CDC N1 and N2 assays and had poor amplification efficiencies (Graber et al., 2021, Wurtzer et al., 2021, Yaniv et al., 2021). A number of biomedical companies have also released products for the detection of SARS-CoV-2 VOCs in wastewater, but have not divulged additional details on their methods. Here we report the development and validation of a panel of AS RT-qPCR assays, which we believe would greatly benefit researchers in both WBE and clinical diagnostics.

There are several important challenges and limitations to consider during interpretation of datasets obtained by the AS-qPCR assays we have described here. First, even though this assay has the potential of tracking the spread and presence of B.1.1.7 on the community level, it remains to be determined if these could be extrapolated to incidence of infection with B.1.1.7 and non-B.1.1.7 strains. For example, differences in virus shedding rates between patients carrying B.1.1.7 versus other strains have not yet been robustly determined, though early evidence does suggest that excretion rates may be higher for the VOC (Kissler et al., 2021). Second, the assays presented here allow for the detection of B.1.1.7 by targeting three mutations. While these are currently highly and solely indicative of the B.1.1.7 VOC, there could be the possibility of detecting emerging strains that may possess the same mutations while constituting a VOC of their own. Nonetheless, the proposed AS RT-qPCR assays can be quickly modified and updated to follow the evolution of B.1.1.7 sequences or other VOCs. Finally, the highly specific and targeted nature of this AS RT-qPCR approach does not allow for its usage to discover new variants. However, the presented method allows for a synergy between clinical genomic surveillance and WBE, where new VOCs can be identified locally via clinical surveillance, followed up with rapid development of AS RT-qPCR assays to target VOC-specific mutations, and finally adopted broadly into WBE campaigns to quickly start monitoring their spread.

Together, this work demonstrates a robust and highly adaptable AS RT-qPCR approach for the detection and genotype discrimination of the SARS-CoV-2 B.1.1.7 variant of concern in wastewater. The presented protocol sets provide a framework for future development of AS RT-qPCR assays to monitor emerging additional variants of concern.

## MATERIALS AND METHODS

### Design of probes and allele specific primers

Primers and probes were designed using Integrated DNA Technologies (IDT)’s PrimerQuest Tool, with the mutation being placed at the 3’ end of either the forward or reverse primer. At least one primer and probe set was designed for each allele-specific (AS) primer direction. All primers were designed to have a Tm in the range of 59–65°C and the probes—in the range of 64–72°C. The probes were designed to anneal to the same strand as the AS primer, with the probe as close to the 3′-end of the AS primers as possible, while avoiding guanines at the 5′-end of the probe. AS primers targeting mutations were designed to include an artificial mismatch near the 3’ terminal nucleotide to increase discrimination between WT and mutant sequences. Primer and probe sequences were analyzed for hairpin, self-dimerization, and heterodimerization. All primers and probes were purchased from IDT. Sequences are shown in **Table 2**.

**Table 2.**
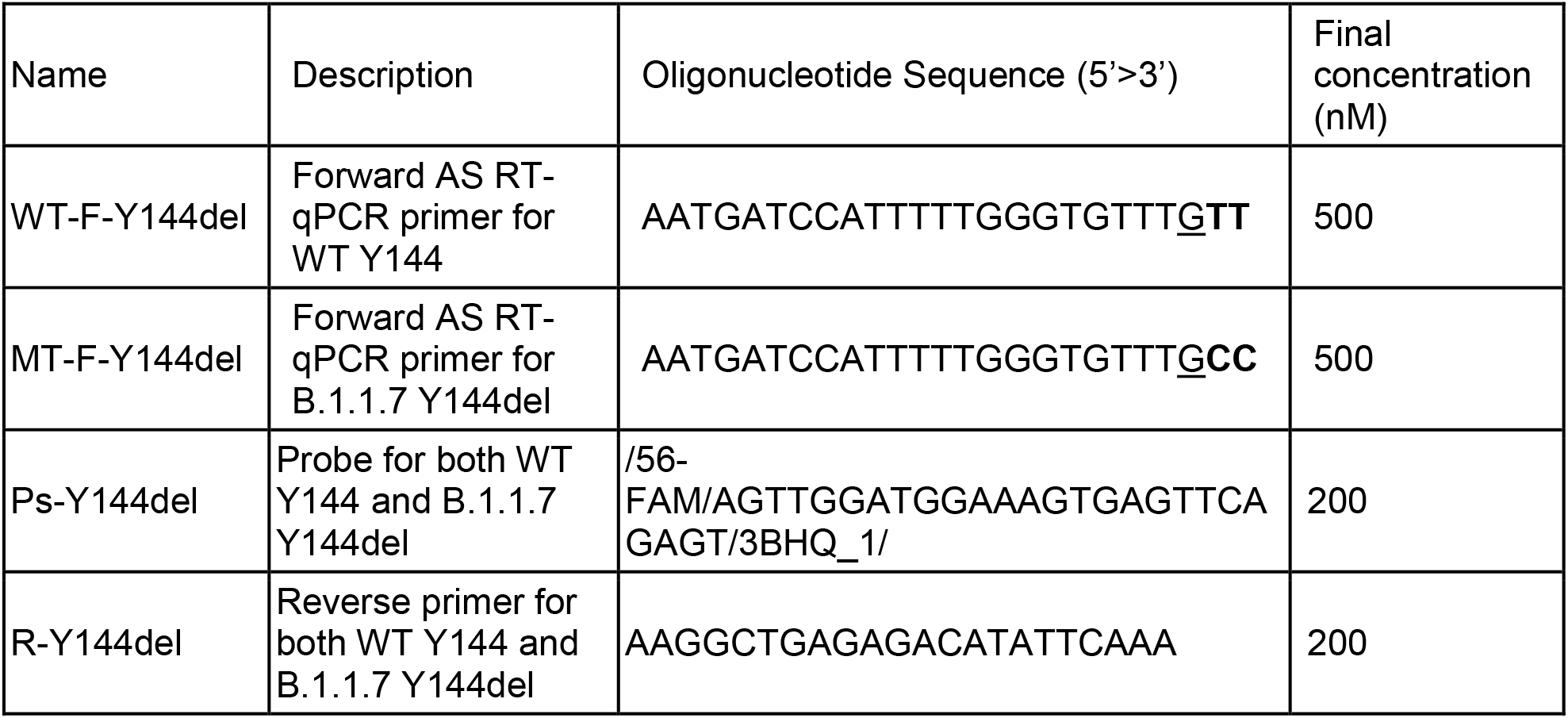
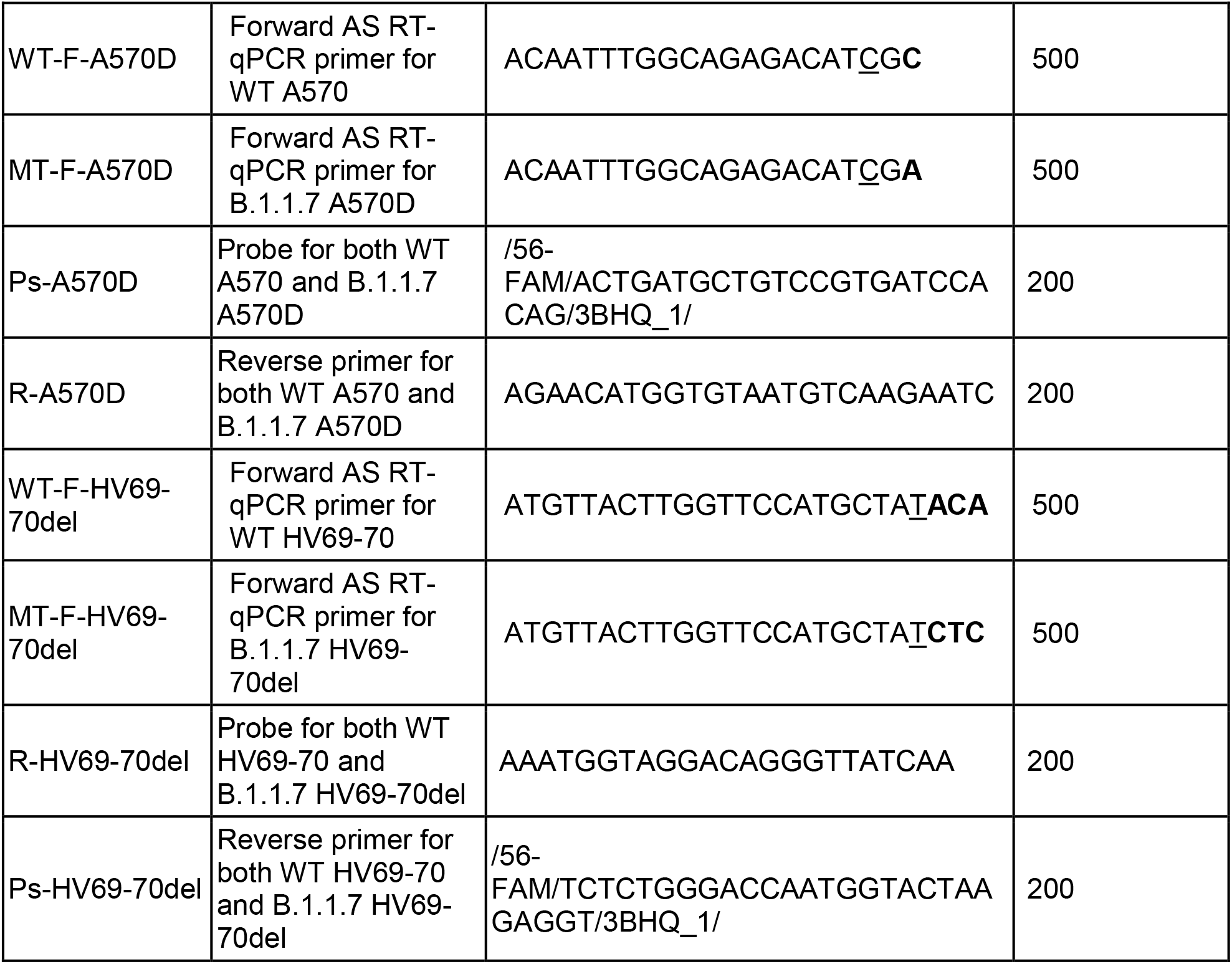
AS RT-qPCR primers and probes sequences developed in this paper. Allele-specific nucleotides are marked bold. Synthetic mismatches are underlined.

### Determination of frequencies of targeted mutations in B.1.1.7 and non B.1.1.7 strains

A total of 732,288 SARS-CoV-2 sequences were downloaded on GISAID (11th March 2021). All the sequences less than 27,000 bp in length or with more than 3,000 N unknown bases are omitted from the analysis. A total of 449,382 sequences remained after QC pre-filtering. 19.8% sequences were assigned as B.1.1.7 strains and 80.2% were assigned as non-B.1.1.7 strains. All the sequences were aligned to the SARS-CoV-2 reference genome (NC_045512.2) using mafft v7.467 to detect the differences of three target mutations in SARS-CoV-2 spike protein (Katoh & Standley, 2013).

### RNA standards

Twist Synthetic SARS-CoV-2 RNA Controls, controls 2 or 4 (Wuhan-Hu-1 or USA/TX1/2020) and control 14 (England/205041766/2020) were used as RNA standards representing WT and B.1.1.7 respectively. RNA standards were prepared as single use aliquots.

### Analysis of AS RT-qPCR primer and probe sets

AS RT-qPCR was performed using the Taqman Virus 1-Step master mix (Thermofisher #4444434) in duplicates, at a final volume of 10 µL, according to the manufacturer’s recommendations. A single reverse or forward primer and probe was used with each set of allele-specific forward or reverse primers (setup is shown in **Table 3**). The RNA templates were analysed at tenfold dilutions, from 10^5^ to 10^1^. The final concentration of the AS RT-qPCR primers were 500 nM, probe at 200 nM, with 1 µL of template. US CDC 2019-nCoV_N1 and 2019-nCoV_N2 (IDT) (sequences shown in **Table 4**) were used following recommendations, at final concentrations of 500 nM primers and 125 nM probe. No template controls were included for each assay and none of them amplified. The reactions are setup using electronic pipettes (Eppendorf) and performed on a Bio-Rad CFX384 real-time PCR instrument under the following conditions, 5 min at 50 °C and 20 s at 95 °C, followed by 45 cycles of 3 s at 95 °C and 30 s at 60 °C.

**Table 3.** The AS RT-qPCR panel for qualitative and quantitative identification of B.1.1.7 and non-B.1.1.7 is made up of six individual RT-qPCR assays.

| Description | Forward primer | Reverse primer | Probe | Amplicon length (bp) |
| --- | --- | --- | --- | --- |
| Detection of HV69-70 | WT-F-HV69-70del | R-HV69-70del | Ps-HV69-70del | 77 |
| Detection of HV69-70del | MT-F-HV69-70del | R-HV69-70del | Ps-HV69-70del | 71 |
| Detection of Y144 | WT-F-Y144 | R-Y144del | Ps-Y144del | 115 |
| Detection of Y144del | MT-F-Y144del | R-Y144del | Ps-Y144del | 112 |
| Detection of A570 | WT-F-A570 | R-A570D | Ps-A570D | 85 |
| Detection of A570D | MT-F-A570D | R-A570D | Ps-A570D | 85 |

**Table 4.**
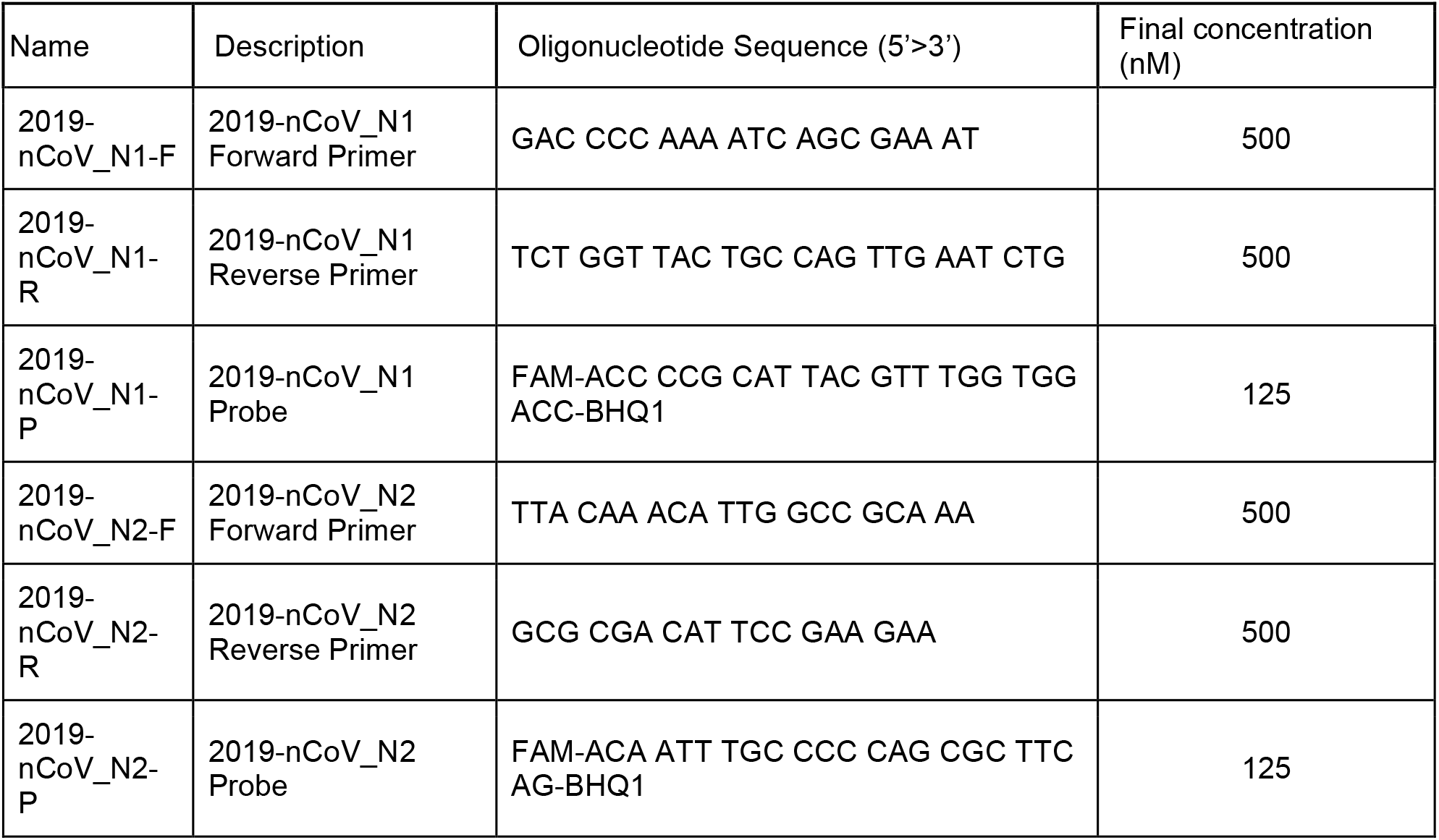
US CDC N gene assay primers and probe sequences.

### Analysis of wastewater samples using AS RT-qPCR panel

24-hour composite samples of raw sewage were obtained from the wastewater treatment plants and selected residential buildings across the US as part of a regular wastewater surveillance service provided by Biobot Analytics, Inc. Samples were pasteurized at 60°C for 1 hour, vacuum filtered through a 0.2 µm membrane (Millipore Sigma) and then sealed and stored at 4°C. 15 ml of clarified samples were concentrated with 10 kDa Amicon Ultra Centrifugal Filter units (Sigma, Cat# UFC9010) to 150 ∼ 200 ul. Samples were then lysed with buffer AVL containing carrier RNA (Qiagen). After adjusting binding conditions with ethanol, samples were subjected to RNA extraction (Qiagen RNeasy kit, Cat# 74182). One-step RT-qPCR was performed as described above, using 1 µL of RNA in a 10 µL reaction. Ct values were converted to RNA copies based on standard curves established with Twist Synthetic SARS-CoV-2 RNA Controls as described above. N1/N2 qPCR quantification was performed as described in Wu et al. 2020.

### Data analysis

Data was analysed using Microsoft Excel and Graphpad prism. Linear regression was performed using Graphpad Prism. Graphs were presented using Graphpad Prism. Results from wastewater samples were analyzed and visualized using Python v3.9.

## Data Availability

Source data will be made available upon request

## Acknowledgements

We thank the sampling operators and treatment plant facilities for their efforts in collecting the wastewater samples, and members of the Biobot Analytics, Inc team for helpful discussions and logistical support. We also thank Stefan Wuertz for helpful discussions and Claire Lim Jiayi and Karl Lars John Berge for help with experiments.

## Contributions

EJA and JT conceptualized the project. WLL and KAM designed the experiments. WLL, KAM, XG, MI, FA, CD, FC, HC, ML, FW, AX, KM analyzed the data. EJA, JT, NG, MM supervised the project. WLL, KAM, XG, FA, FC, SM, RF, MMP, and STW performed experiments. All authors contributed to writing the manuscript. All authors read and approved the manuscript.

